# Increased Risks of Large-Artery Atherosclerosis and Brainstem Stroke in Patients With Nasopharyngeal Carcinoma

**DOI:** 10.1101/2023.10.23.23297445

**Authors:** Chin-Yang Lin, Cheng-Li Lin, Ji-An Liang, Yao-Ching Wang, Ying-Chun Lin, Chih-Ying Liao, Ting-Chun Lin

## Abstract

**Background:** Information about the characteristics of stroke is crucial for improved screening, prevention, and treatment. The pattern of ischemic stroke in patients with nasopharyngeal carcinoma (NPC) is not well understood. The objective of this study is to investigate the etiology of ischemic stroke and the affected vascular territories in patients with NPC.

**Methods:** The Taiwan National Health Insurance Research Database and the Taiwan Stroke Registry were combined to identify the following 2 cohorts: patients with NPC having a subsequent stroke (NPC cohort) and patients without NPC having a stroke (non-NPC cohort). Data pertaining to the period from 2006 to 2018 were analyzed. The TOAST (Trial of ORG 10 172 in Acute Stroke Treatment) subtype of ischemic stroke and the affected vascular territories were analyzed and compared between the 2 cohorts.

**Results:** The NPC cohort comprised 277 patients, whereas the non-NPC cohort comprised 125 024 patients. The most common TOAST subtype of ischemic stroke was large-artery atherosclerosis (LAA) in the NPC cohort and small-vessel occlusion in the non-NPC cohort. The prevalence of LAA was considerably higher in the NPC cohort (41.9%) than in the non-NPC cohort (28.3%). The analysis of the affected vascular territories revealed that the incidence of brainstem stroke was significantly higher in the NPC cohort (25.4%) than in the non-NPC cohort (14.5%).

**Conclusions:** Patients with NPC tend to have increased risks of LAA and brainstem ischemic stroke. Physicians should consider sparing the vertebral and basilar arteries during definitive radiotherapy for NPC. Long-term follow-up for patients with NPC may include brain magnetic resonance imaging. This study may guide radiation oncologists in clinical decision-making.

## Introduction

Nasopharyngeal carcinoma (NPC) is a prevalent cancer in Asian countries.^1^ In Taiwan, more than 1000 new cases of NPC are reported annually.^2^ The standard treatment for NPC is radiotherapy (RT) for stage I disease and concurrent chemotherapy and RT for all other stages. The radiation field consistently involves the nasopharynx (dose, approximately 70 Gy, including the lymph nodes) and bilateral neck (dose, 50 to 60 Gy).^3^ Radiotherapy to the neck can cause ischemic stroke, even after several years.^3-5^

The main cause of RT-induced stroke is generally presumed to be post-RT vasculopathy, which leads to carotid artery stenosis and plaque formation.^7,8^ Attempts have been made to reduce the risk of RT-induced ischemic stroke with techniques such as carotid artery–sparing or vertebral artery–sparing intensity-modulated RT and proton therapy, but the effectiveness of these techniques remains unclear.^9,10^

The Trial of ORG 10 172 in Acute Stroke Treatment (TOAST) classification divides ischemic stroke etiology into 5 subtypes: large-artery atherosclerosis (LAA), small-vessel occlusion (SVO), cardioembolism, stroke of other determined etiology, and stroke of an undetermined cause.^11,12^ In addition to standard risk management, determining the precise subtype of stroke is crucial for the prognosis and further long-term treatment–related decision-making.^13,14^ Strokes can also be divided according to the affected vascular territory into anterior circulation strokes (mainly supplied by the internal carotid artery) and posterior circulation strokes (supplied by the basilar and vertebral arteries).^1,2^ In the general population, the middle cerebral artery of the anterior circulation is the most commonly affected territory in ischemic stroke.^15^ However, in patients with NPC, the distribution of posttreatment stroke subtypes according to their location or etiology remains unclear.

In this study, we combined Taiwan’s National Health Insurance Research Database (NHIRD) and the Taiwan Stroke Registry (TSR), a multicenter stroke registry database, to analyze nationwide real-world data and identify the characteristics of subsequent ischemic stroke in patients with NPC, including its common TOAST subtypes and affected vascular territories.

## Methods

### Databases

For this real-world nationwide cohort study, we linked the TSR and NHIRD. The NHIRD contains detailed registry and claim data of all residents of Taiwan who are covered under Taiwan’s mandatory National Health Insurance program.^16^ The TSR is sponsored by Taiwan’s Department of Health and Welfare and has a nationwide network of stroke centers. It contains the records of the stroke types (e.g., ischemic stroke, transient ischemic attack, intracerebral hemorrhage, subarachnoid hemorrhage, and other), diagnostic examinations (e.g., computed tomography [CT] and magnetic resonance imaging [MRI]), and the 5 major subtypes of ischemic strokes according to the TOAST criteria (LAA, SVO, cardioembolism, specific pathogenesis, and undetermined pathogenesis).^17,18^

## Results

### Study Population and Cohort Design

From the TSR, we identified patients who had a stroke between August 1, 2006, and August 31, 2018. Next, patients with a previous diagnosis of NPC (*International Classification of Diseases*, *Ninth Revision*, *Clinical Modification* code: 147; *International Classification of Diseases*, *Tenth Revision*, *Clinical Modification* code: C11) were identified from the NHIRD.^16,19^ The NHIRD and TSR were linked using encrypted and unique personal identification numbers. Thus, we identified a cohort of patients with stroke who had a history of NPC. In addition, we established a comparison cohort comprising patients with stroke without NPC (**Figure 1**). All included patients were aged ≥18 years.

**Figure 1.**
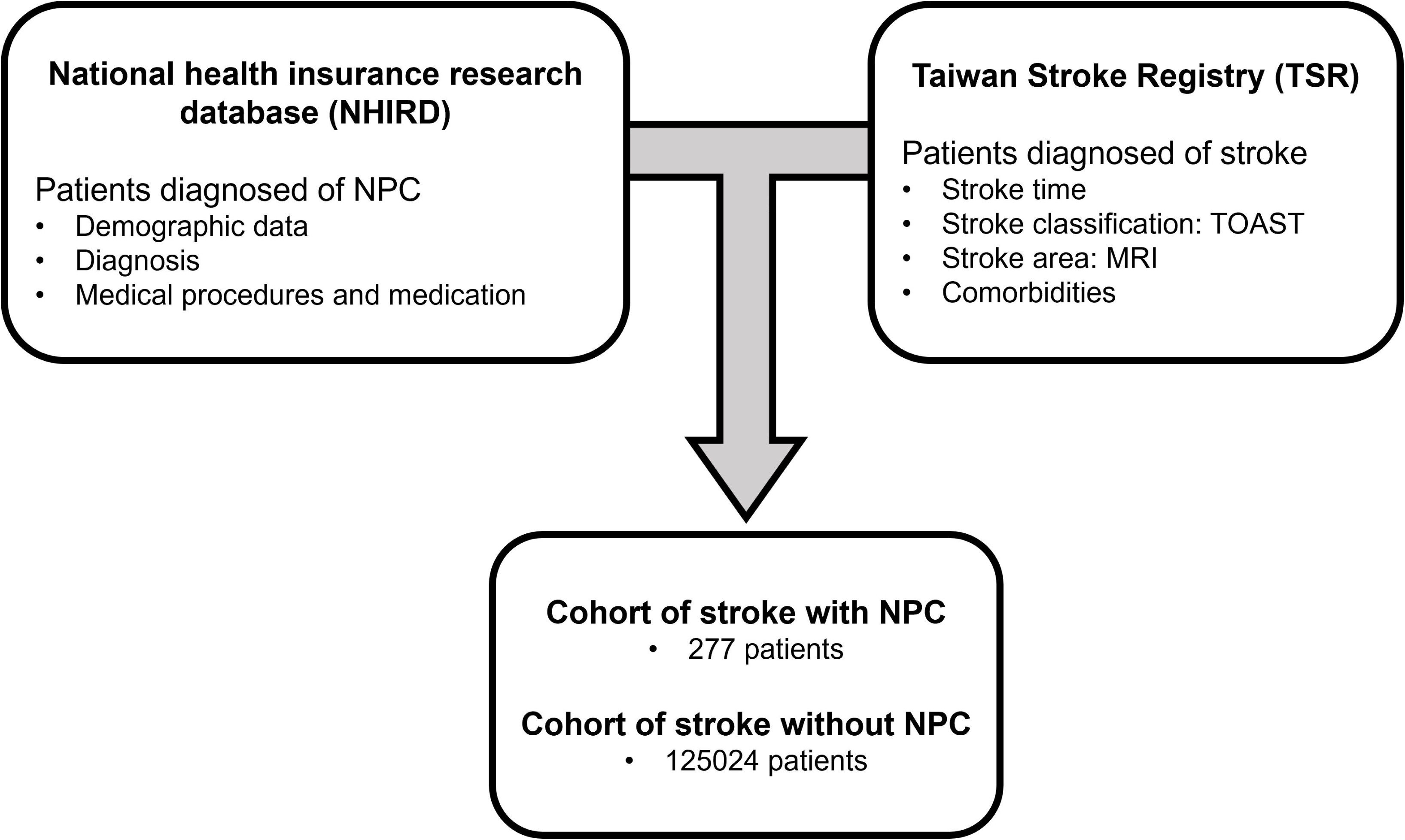
Flow diagram of cohort identification.

### Ethical Approval

This study was approved by the local Institutional Review Board.

### Analysis of the Vascular Territories Affected by Ischemic Stroke

To accurately define the stroke area, we used MRI data to identify the vascular territories affected by ischemic stroke (available in the TSR). These territories were broadly classified into anterior and posterior circulation groups. The anterior circulation group comprised the ischemic stroke–affected anterior and middle cerebral arteries, whereas the posterior circulation group comprised the ischemic stroke–affected posterior cerebral artery (PCA), thalamus, brainstem, and cerebellum.

### Statistical Analysis

Descriptive data are presented in terms of the frequency (percentage) or the mean ± standard deviation values. The NPC and non-NPC cohorts were compared in terms of sex, age, stroke comorbidities, and cancer treatment by using the chi-square or independent *t* test. We analyzed the following stroke-related comorbidities: type 2 diabetes mellitus, hypertension, hyperlipidemia, chronic obstructive pulmonary disease, chronic kidney disease, coronary artery disease, and atrial fibrillation. Statistical significance was set at *P* < .05. All statistical analyses were performed using SAS (version 9.4; SAS Institute, Cary, NC, USA).

### Cohort Characteristics and Stroke Types

The NPC cohort had more men (80.9% vs 60.2%), younger patients (61.7 ± 11.2 vs 67.5 ± 13.7), and fewer comorbidities (hyperlipidemia, chronic obstructive pulmonary disease, coronary artery disease, and atrial fibrillation) than did the non-NPC cohort. In the NPC cohort, 90.6% received RT and 73.3% received chemotherapy (**Table 1**). The average time interval between NPC diagnosis and stroke was 6.9 years (median: 6.4; interquartile range: 3.8–9.7).

**Table 1.** Clinicodemographic Characteristics of the Patients.

| Variables | Stroke With NPC |  | Stroke |  | P-Value |
| --- | --- | --- | --- | --- | --- |
|  | History |  | Without NPC |  |  |
|  | N=277 |  | N=125024 |  |  |
|  | n | (%) | n | (%) |  |
| Gender |  |  |  |  | <0.001 |
| Female | 53 | 19.1 | 49806 | 39.8 |  |
| Male | 224 | 80.9 | 75218 | 60.2 |  |
| Age group (years) |  |  |  |  | <0.001 |
| 20-49 | 36 | 13 | 14598 | 11.7 |  |
| 50-64 | 146 | 52.7 | 38201 | 30.6 |  |
| >=65 | 95 | 34.3 | 72225 | 57.8 |  |
| Age (years), mean ± standard deviation (range) | 61.7±11.2 (33-88) |  | 67.5±13.7 (18-109) |  | <0.001 |
| Comorbidities |  |  |  |  |  |
| Type 2 DM | 30 | 10.8 | 9722 | 7.78 | 0.06 |
| Hypertension | 207 | 74.7 | 98780 | 79 | 0.08 |
| Hyperlipidemia | 118 | 42.6 | 61984 | 49.6 | 0.02 |
| COPD | 62 | 22.4 | 34946 | 28 | 0.04 |
| CKD | 17 | 6.14 | 6450 | 5.16 | 0.46 |
| CAD | 76 | 27.4 | 52441 | 41.9 | <0.001 |
| AF | 9 | 3.25 | 9243 | 7.39 | 0.008 |
| Treatment |  |  |  |  |  |
| Chemotherapy | 203 | 73.3 | 2499 | 2 | <0.001 |
| Radiotherapy | 251 | 90.6 | 16 | 0.01 | *<0.001* |

DM: Diabetes mellitus; COPD: Chronic obstructive pulmonary disease; CKD: Chronic
kidney disease; CAD: Coronary artery disease; AF: Atrial fibrillation.

In both cohorts, the predominant type of stroke was ischemic stroke (77.6% and 72.7%, respectively), followed by intracerebral hemorrhage, transient ischemic attack, and subarachnoid hemorrhage (**Table 2**). The distribution of stroke types was comparable between the 2 cohorts.

**Table 2.** Types of Stroke in Patients With or Without Nasopharyngeal Carcinoma.

| Variable | Stroke With NPC History |  | Stroke Without NPC |  | P-Value |
| --- | --- | --- | --- | --- | --- |
|  | N=277 |  | N=125024 |  |  |
|  | n | (%) | n | (%) |  |
| Stroke type |  |  |  |  | 0.30 |
| Ischemic stroke | 215 | 77.6 | 90830 | 72.7 |  |
| TIA | 15 | 5.42 | 8582 | 6.86 |  |
| ICH | 34 | 12.3 | 17925 | 14.3 |  |
| SAH | 3 | 1.08 | 3384 | 2.71 |  |
| Other | 10 | 3.61 | 4303 | 3.44 |  |

TIA: Transient ischemic attack; ICH: Intracerebral hemorrhage; SAH: Subarachnoid hemorrhage.

### Between-Group Differences in the TOAST Subtypes of Ischemic Stroke

In the NPC cohort, 215 patients had ischemic stroke (**Table 2**); of them, TOAST subtype was documented for 186 patients; the main subtype was LAA (41.9%), followed by SVO (25.3%), undetermined cause (21.5%), other determined etiology (5.9%), and cardioembolism (5.4%). This distribution varied significantly from that in the non-NPC cohort: SVO (39.0%), LAA (28.3%), undetermined cause (18.3%), cardioembolism (12.7%), and other determined etiology (1.8%; **Table 3**).

**Table 3.** TOAST Classification for Ischemic Stroke in Patients With or Without Nasopharyngeal Carcinoma.

| Variable | Stroke With NPC History |  | Stroke Without NPC |  | P-Value |
| --- | --- | --- | --- | --- | --- |
|  | N=186 |  | N=84752 |  |  |
|  | n | % | n | % |  |
| TOAST classification |  |  |  |  | <0.001 |
| Large artery<br>atherosclerosis (LAA) | 78 | 41.9 | 23954 | 28.3 |  |
| Small vessel<br>occlusion (SVO) | 47 | 25.3 | 33067 | 39.0 |  |
| Cardioembolism | 10 | 5.4 | 10723 | 12.7 |  |
| Specific etiology | 11 | 5.9 | 1497 | 1.8 |  |
| Undetermined etiology | 40 | 21.5 | 15511 | 18.3 |  |

### Between-Group Differences in the Vascular Territories Affected by Ischemic Stroke

MRI data were available for 138 patients with NPC (49.8%) and 57 711 patients without NPC (46.2%). Regarding the affected vascular territories, the incidence rates of anterior circulation stroke were similar between the 2 cohorts but that of posterior circulation stroke was nonsignificantly higher in the NPC cohort than in the non-NPC cohort (31.2% vs 24.4%, respectively; *P* = .07). Notably, the incidence rate of brainstem stroke was significantly higher in the NPC cohort than in the non-NPC cohort (25.4% vs 14.5%, respectively; *P* < .001; **Table 4**).

**Table 4.**
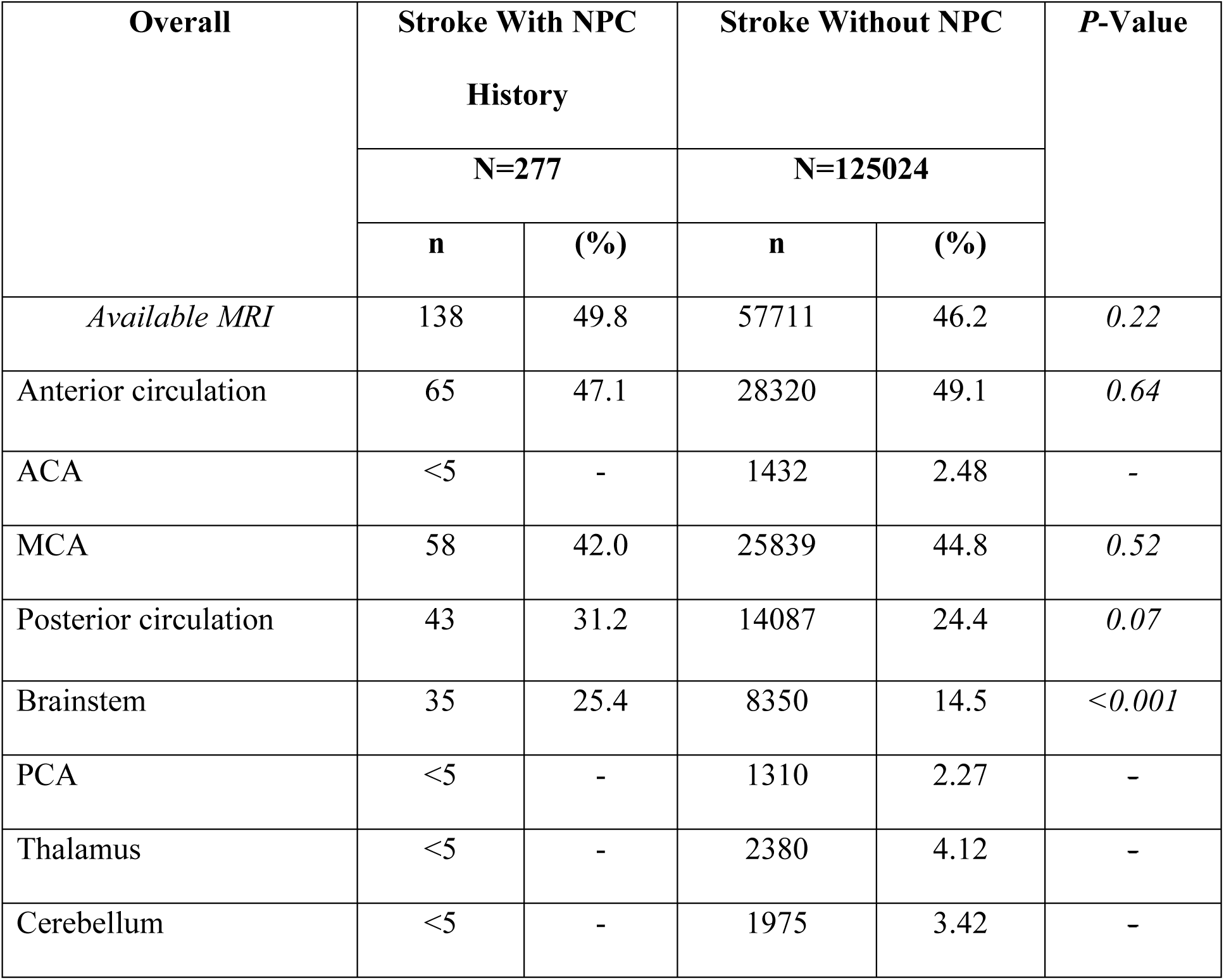
Anatomical Distribution of Ischemic Stroke in Patients With or Without Nasopharyngeal Carcinoma.

## Discussion

Patients undergoing RT have an elevated risk of ischemic stroke.^20^ In Taiwan, NPC is a common cancer, particularly in men; it has been associated with smoking and Epstein–Barr virus infection.^2,21,22^ Patients with NPC often have long survival, and some of them live long enough to develop ischemic stroke many years after receiving definitive concurrent chemotherapy and RT.^6,23,24^ Studies have indicated RT as the main cause of ischemic stroke in patients with head and neck cancer, with late complications including vasculopathy and fibrosis as the primary underlying mechanisms.^5,25^ Although researchers have frequently discussed why ischemic stroke develops after RT, few have characterized ischemic stroke, such as in terms of the TOAST classification or anatomically on the basis of affected vascular territories, in patients receiving RT.

Our results revealed 2 distinct patterns of ischemic stroke subsequent to an NPC diagnosis. First, the etiology of ischemic stroke was predominantly LAA in patients with NPC compared with SVO in the general population without NPC; the prevalence of LAA was approximately 1.5 times higher in the NPC cohort than in the non-NPC cohort (41.9% vs 28.3%, respectively). Second, the posterior circulation arteries were more susceptible to vascular injuries in the NPC cohort than in the non-NPC cohort. The incidence rate of brainstem stroke was approximately 1.75 times higher in the NPC cohort than in the non-NPC cohort (25.4% vs 14.5%, respectively). Together, these results suggest that the increase in the incidence of stroke in the NPC cohort results, at least partially, from atherosclerosis in large posterior circulation arteries, particularly the basilar and vertebral arteries.

No high-level evidence is available to explain how stroke is developed after RT. This is challenging to determine through in vivo or in vitro studies not only because radiation-induced late effect takes years to manifest but also because this effect is an interaction between the vessels and the microenvironment, which is difficult to emulate in a laboratory. Several case reports have characterized radiation-induced vasculopathy as concentric vessel wall thickening resulting from chronic vasculitis.^25-27^ We noted a higher prevalence of LAA in patients with NPC than in individuals without NPC; our findings align with the mechanisms proposed by the aforementioned studies.

The differences in the anatomical characteristics between the carotid arteries (supplying the anterior circulation) and the posterior circulation arteries that may be relevant in the context of the present study are as follows. First, the brainstem area has no well-established collateral circulation. For the internal carotid artery, the circle of Willis at least partially compensates for the insufficient blood flow. The cerebellum has collateral flow between different cerebellar arteries, including superior cerebellar arteries, anterior inferior cerebellar artery, and posterior inferior cerebellar artery. The PCA and thalamus may also receive blood supplies from the anterior circulation via the posterior communicating arteries. Nonetheless, brainstem perforators directly branch from the basilar artery. Therefore, pathological changes in the basilar artery may compromise the bifurcation points, resulting in brainstem stroke. Second, the diameter of the brainstem perforators is substantially smaller (80 to 840 µm) than that of the internal carotid artery (ICA; 4 to 6 mm).^28-30^ This may also explain why the posterior circulation may be more susceptible to atherosclerosis-related vascular occlusion. Third, early identification of pathological changes, such as plaque formation and stenosis, in the posterior circulation may be challenging because the cranial and vertebral bony structures limit their visualization on transcranial Doppler ultrasound. Thus, obtaining detailed information through extracranial carotid Doppler ultrasound is more challenging for the posterior circulation than for the anterior circulation. The brainstem and thalamus are usually the blind spots in CT and MRI perfusion examinations. Finally, in curative RT for NPC, high-dose radiation (usually ≥70 Gy) is conventionally delivered to the clivus and skull base, either intentionally or unintentionally, because of their close proximity to the tumor at the nasopharynx. The basilar and vertebral arteries are usually impossible to spare, which results in full-dose and full-length irradiation of the arteries supplying the posterior circulation. By contrast, only a partial length of the ICA (a considerably long and thick artery) receives a partial dose (usually 40 to 70 Gy). Higher doses of RT may also lead to relatively higher injury in the arteries of the posterior circulation.

Clinically, our findings may guide radiation oncologists in making decisions regarding treatment and follow-up for patients with NPC. In particular, they may consider contouring the basilar and vertebral arteries noted on the MRI during treatment planning, identifying them as at-risk arteries. Although the optimal artery dose constraint for averting vascular injuries remains unknown, proton therapy may be preferred over photon therapy when prioritizing artery sparing.^10^ Regarding follow-up, patients with NPC commonly undergo annual carotid echocardiography, but vascular changes in the brain are usually overlooked.^3^ We suggest performing brain MRI 7 years after RT to analyze the risk of posterior circulation ischemic stroke. In addition, these patients should be referred to specialists for the management of blood pressure. LAA is typically treated through antithrombotic therapy, stenting, and endarterectomy.^14^ Thus, for long-term NPC survivors after definitive RT, baseline ultrasonography screening, more frequent follow-up visits, and perhaps a reduced threshold for stents or similar interventions can be considered.

The strengths of our study include the use of an ideal cohort by selecting NPC as the target population. These patients are usually relatively young, have a good performance status, are long-term survivors, and receive consistent treatment. A nationwide study conducted in Taiwan in 2015 revealed that the rates of 5-year overall survival in patients with any-stage NPC and those with early-stage NPC were 65.2% and 90%, respectively; these data enabled us to observe the onset of subsequent stroke.^2,32,33^ Had we selected a cancer type with worse survival, we might not have been able to observe the development of this stroke. Additionally, patients with NPC usually receive RT with a consistent field and chemotherapy with a consistent regimen; the RT field covers the nasopharynx, skull base, and bilateral neck, and the chemotherapy regimen is concurrent cisplatin-based chemotherapy. Thus, the radiation volume and dose to the carotid and basilar arteries as well as the chemotherapy regimen and schedule do not vary markedly across patients with NPC receiving definitive treatment.

This study has several limitations, mainly resulting from its retrospective and registry-based nature. First, although the NHIRD did not record the detailed RT field and dose and did not always include the American Joint Committee on Cancer staging for each patient. Thus, some patients with NPC might have received local RT with a palliative intent, such as pain or symptom control. Nonetheless, this limitation likely does not affect our findings because patients receiving palliative RT were unlikely to survive long enough to develop ischemic stroke. Second, the RT technique applied in the time range of this cohort probably included older modalities, such as two- or three-dimensional conformal RT, that are not extensively used in modern-day Taiwan. In the era of intensity-modulated RT, fewer late complications, such as post-RT stroke, may occur in patients with NPC. Finally, the NPC cohort had insufficient number of patients with available MRI data, precluding the comprehensive analysis of the stroke subgroups. We could only conclude whether the incidence of anterior cerebral artery, PCA, thalamus, and cerebellum stroke was considerably lower in the NPC cohort than in the non-NPC cohort. Moreover, the TSR data did not record the specific vessels that were stenotic or occluded.

Despite these limitations, given the prolonged interval between RT and ischemic stroke incidence in patients with NPC, the onset of a stroke is difficult to observe in a clinical trial setting. Thus, this retrospective, registry-based study might have been the best approach for identifying the characteristics of posttreatment ischemic stroke in patients with NPC. Our findings provide insights into the characteristics and possible etiology of posttreatment ischemic stroke in patients with a history of NPC.

## Data Availability

Research data will be shared upon request to the corresponding author.

## Acknowledgments

TCL and CYL designed the study and drafted the manuscript. CLL collected and analyzed the data. JAL, YCW, YCL and CYL reviewed and edited the manuscript. We also thank Doctor Chun-Ru Chien and Wallace Academic Editing for proofreading this manuscript. All authors contributed to the article and approved the submitted version.

## Sources of Funding

None.

## Disclosures

Authors have no conflict of interest to declare.

## Notes

### Competing Interest Statement

The authors have declared no competing interest.

### Funding Statement

No funding was received for conducting this study.

### Author Declarations

This study was approved by the Institutional Review Board of China Medical University Hospital. Approval number: CMUH109-REC2-031(CR-3).

